# Brief Report-Combination Capmatinib and Trametinib in Metastatic MET-driven Non-Small Cell Lung Cancer

**DOI:** 10.64898/2026.05.19.26353265

**Authors:** Tyiesha Brown, Matthew S. Lara, Fei Jiang, Edward B. Garon, Jonathan W. Goldman, Jonathan W. Riess, Collin M. Blakely

## Abstract

**Introduction:** MET tyrosine kinase (TKI) therapy has improved outcomes in patients with non-small cell lung cancer (NSCLC) harboring MET alterations. However, primary and acquired resistance ultimately limits durability of response. This study evaluated the safety and efficacy of the MET inhibitor capmatinib with the MEK inhibitor trametinib in patients with metastatic MET-driven NSCLC who had progressed on prior treatment with at least one MET inhibitor.

**Methods:** A multicenter phase I study evaluated capmatinib in combination with trametinib in patients with advanced stage NSCLC harboring activating MET alterations and prior exposure to at least one MET TKI. A 3+3 dose-escalation design was employed to assess safety and tolerability of the combination.

**Results:** Three patients (n = 3) were enrolled in the study and completed a median of 3 cycles of therapy. Dose-limiting toxicities, including rash, edema, and nausea, necessitated dose reductions in the first two patients and initiation of the third patient at a lower dose level. Ultimately, all patients discontinued therapy due to treatment-related adverse events. The study was terminated early due to poor accrual and TRAEs. No radiographic objective responses were observed.

**Conclusions:** In this phase I trial, capmatinib plus trametinib was associated with significant treatment-related adverse events and treatment was discontinued in all participants. Based on these findings, further investigation of this combination of MET and MEK inhibitors is not recommended.

## Introduction

MET alterations occur in approximately 2–4% of NSCLC cases and are most frequently observed in lung adenocarcinoma. MET encodes a receptor tyrosine kinase that, upon binding its ligand, hepatocyte growth factor (HGF), undergoes dimerization and activates multiple downstream signaling pathways (such as RAS/MAPK, PI3K/AKT) critical for tumor growth, survival, and invasion (1).

Targeted inhibition of MET with the tyrosine kinase inhibitors (TKIs), capmatinib and tepotinib, has improved outcomes for patients with advanced NSCLC harboring MET alterations, as demonstrated in clinical observations and the GEOMETRY and VISION trials (2-3, 6-14). Despite these advances, resistance to MET TKI therapy invariably occurs. Through comprehensive sequencing of circulating tumor DNA from 289 patients with MET exon 14 (METex14) mutated NSCLC, we revealed a significantly higher prevalence of co-occurring alterations in RAS-MAPK pathway genes compared with tumors harboring EGFR mutations. Furthermore, in a preclinical METex14 mutated model, we found that NF1 downregulation or KRAS overexpression was associated with enhanced MAPK signaling and resistance to MET TKI therapy. This resistance was overcome by dual MET and MEK inhibitor therapy, suggesting that a combination therapeutic approach directed towards MET and MAPK pathway signaling warrants clinical investigation (5).

## Methods

This multi-institution, investigator-initiated, phase I/Ib trial funded by Novartis Pharmaceuticals Corporation, evaluated capmatinib plus trametinib in patients with previously treated Stage IV non-small cell lung cancer with MET exon 14 skipping mutation. A 3+3 dose escalation scheme was employed with four prespecified dose levels. Dose level -2 consisted of capmatinib 200 mg twice daily plus trametinib 1.0 mg daily; dose level -1, capmatinib 300 mg twice daily plus trametinib 1.0 mg daily; dose level 1, capmatinib 300 mg twice daily plus trametinib 1.5 mg daily; and dose level 2, capmatinib 400 mg twice daily plus trametinib 1.5 mg daily (**Supplementary Figure 1**). Dose-limiting toxicity (DLT) was defined as any of the following toxicities possibly related to study treatment occurring within the first cycle:

Grade ≥ 3 clinically significant non-hematologic toxicity; Grade ≥ 4 neutropenia or thrombocytopenia lasting > 7 days; Grade ≥ 3 febrile neutropenia; Any adverse event requiring a dose reduction in the DLT period will be considered a DLT; Rash or diarrhea Grade ≥ 3 lasting > 3 days attributed to study treatment despite optimal medical management. The primary endpoint of the study was to determine the safety and tolerability of the combination and determine the recommended phase 2 dose (RP2D). For additional methods, see supplementary appendix.

## Results

A total of three patients were enrolled in this study (**Supplementary Figure 2**). The baseline characteristics of patients treated are summarized in **Table 1**. All three patients never smoked and had stage IV NSCLC with MET exon 14 skipping mutations. Two patients had received one prior line of MET inhibitor therapy, while one patient had two prior lines of systemic therapy including combination chemotherapy and immunotherapy. Two patients initiated treatment at dose level 1. During cycle 1, one patient experienced a grade 2 rash requiring a dose hold of more than 7 days, this was deemed a dose-limiting toxicity (DLT). The second patient did not experience a DLT during the DLT period, however, during cycle 2, the patient’s nausea worsened to grade 2 and edema worsened to grade 3 which required the patient to hold dose for approximately 20 days. The patient restarted treatment at a dose level -1. Although the grade 3 edema requiring dose hold and reduction occurred outside the DLT window, the event was classified as a DLT by the investigator due to its severity and the prolonged dose hold required. Due to the frequency and severity of adverse events observed in the first two patients at the dose level 1, the third patient was initiated at dose level -1. Treatment-related adverse events for the third patient included grade 1 facial rash and serious adverse event of grade 3 pneumonitis, resulting in treatment discontinuation.

**Table 1:** Baseline Characteristics of Patients.

| **Patient Characteristic** | **Number of patients**<br>**(%)** |
| --- | --- |
| Age, median (range) | 75 (64 – 77) |
| Sex |  |
| M | 1 (33) |
| F | 2 (67) |
| History of Smoking |  |
| Current/Former | 0 (0) |
| Never | 3 (100) |
| Prior Lines of systemic therapy |  |
| 1 | 2 (67) |
| 2 | 1 (33) |
| Prior Lines of MET inhibitor therapy |  |
| 1 | 3 (100) |
| MET Exon 14 Skipping | 3 (100) |

The most frequently observed adverse events (AEs) and treatment related adverse events (TRAEs) of any grade are listed in **Table 2 and Supplementary Table 1** and included rash (n = 3; 100%), dyspnea (n = 2; 67%), edema (n = 2; 67%), nausea (n = 2; 67%) and ocular irritation (n = 2; 67%). The most common study attributable grade 3 or higher AEs were edema (n = 2; 67%) and pneumonitis (n = 1; 33%). Two serious adverse events were reported including pneumonitis and chest pain. The median number of treatment cycles was three. All patients discontinued the study due to AEs. There were no treatment related deaths.

**Table 2:** Summary of Adverse Events Observed.

| **Toxicity Observed** | **Grade 1 (%)** | **Grade 2 (%)** | **Grade 3 (%)** | **Total, Any Grade (%)** |
| --- | --- | --- | --- | --- |
| Rash | 2 (67) | 1 (33) |  | 3 (100) |
| Ocular irritation* | 2 (67) |  |  | 2 (67) |
| Dyspnea | 2 (67) |  |  | 2 (67) |
| Edema |  |  | 2 (67) | 2 (67) |
| Nausea | 1 (33) | 1 (33) |  | 2 (67) |
| Vomiting |  | 1 (33) |  | 1(33) |
| Esophagitis |  | 1 (33) |  | 1(33) |
| Pneumonitis |  |  | 1 (33) | 1(33) |
| Rhinorrhea | 1 (33) |  |  | 1(33) |
| Diarrhea | 1 (33) |  |  | 1(33) |
| Vitreous detachment | 1 (33) |  |  | 1(33) |
| Mouth sores | 1 (33) |  |  | 1(33) |
| Chest pain |  |  | 1 (33) | 1(33) |
| Shoulder pain | 1 (33) |  |  | 1(33) |
| Dysuria | 1 (33) |  |  | 1(33) |
| Urinary tract infection |  | 1 (33) |  | 1(33) |
| Melena | 1 (33) |  |  | 1(33) |
| Colon Polyps |  | 1 (33) |  | 1(33) |
| Pruritus | 1 (33) |  |  | 1(33) |

\*Ocular irritation included conjunctival injection, xerophthalmia, and periorbital edema

All patients experienced stable disease (SD) as their best response per RECIST 1.1 criteria. The best percentage change in target lesion size ranged from –20% to +14% (**Figure 1**). No patient experienced disease progression during study treatment. All patients discontinued the study prior to trial closure, due to adverse events. The study was ultimately terminated prior to evaluation of dose level -2 due to poor accrual and treatment-related toxicity. Median progression-free survival, overall survival, and median duration of response could not be estimated.

**Figure 1:**
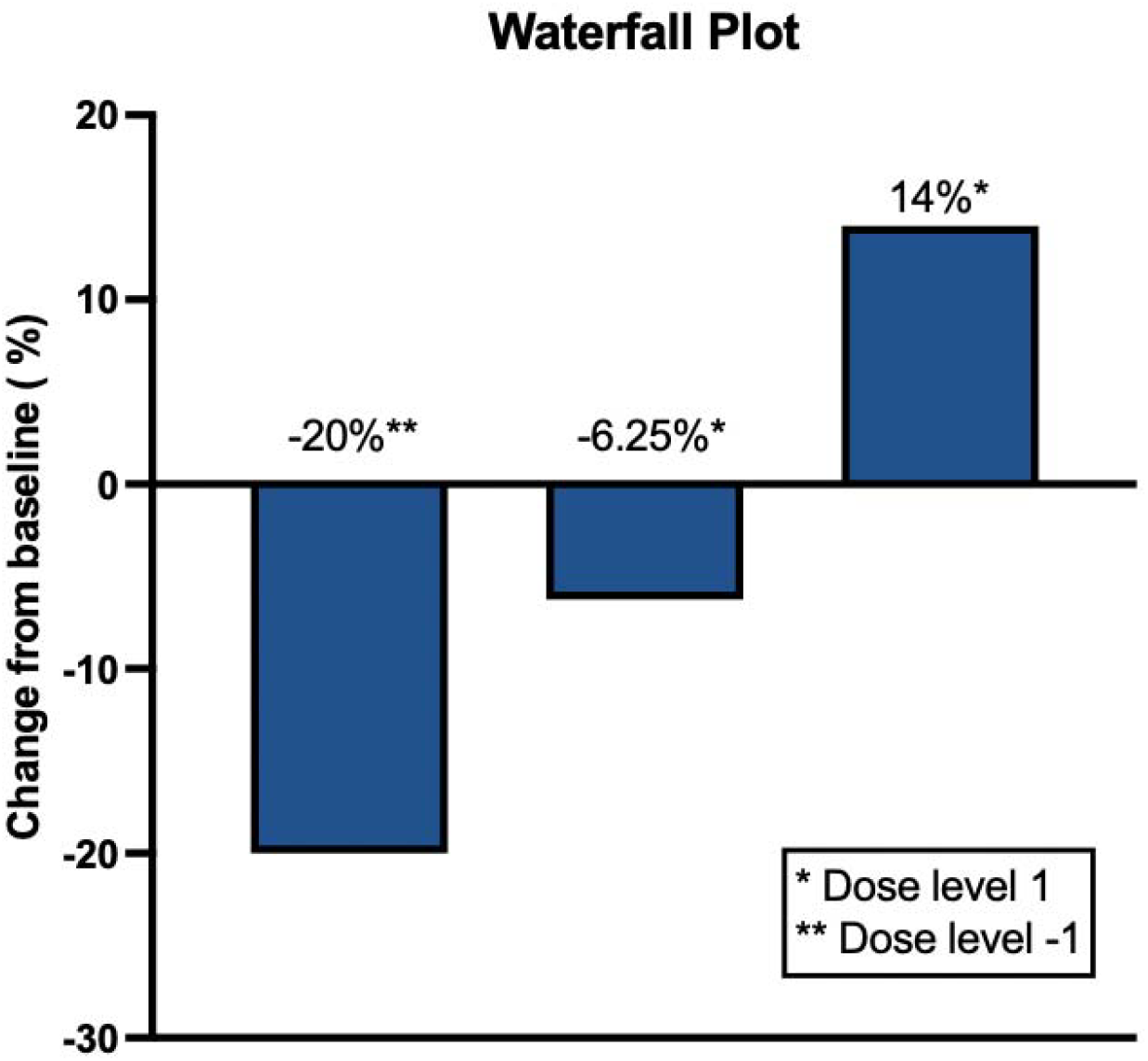
Waterfall Plot for Best Percentage Change, per RECIST criteria.

Best percentage change from baseline in target lesions. Waterfall plots for best percentage change in target lesion size by RECIST 1.1 criteria by investigator assessment are shown for patients who received of capatinib + trametinib. * Indicates patients who received dose level 1 of capmatinib 300 mg PO twice daily plus trametinib 1.5 mg PO daily. ** Indicates patients who received dose level -1 of capatinib 300 mg PO twice daily plus trametinib 1.0 mg PO daily.

## Discussion

This study sought to characterize the safety and efficacy of the combination of the MET inhibitor capmatinib with the MEK-inhibitor trametinib in patients with MET-altered NSCLC who had progressive disease on prior MET inhibitor therapy. The primary finding of this study was the poor tolerability of the drug combination at dose level 1, with capmatinib 300 mg orally twice daily and trametinib 1.5 mg orally once daily. This resulted in dose reductions to dose level -1 using capmatinib 300 mg orally twice daily and trametinib 1.0 mg orally once daily. Although no treatment-related deaths occurred, dose-limiting toxicities and frequent drug-related adverse events precluded dose escalation and ultimately led to treatment discontinuation in all patients. The anticipated adverse events for MEK and MET combination included fatigue, nausea, vomiting, diarrhea, rash, and peripheral edema. The known U.S. Food and Drug Administration (FDA) adverse events for capmatinib monotherapy include pneumonitis (4.5%), peripheral edema (53%), and rash (13%). Trametinib has FDA-reported adverse events of pneumonitis (2%), peripheral edema (21%), and rash (32 %). All reported percentages represent all-grade adverse events. Edema is a common and challenging toxicity to manage in patients treated with MET inhibitors, and in this study, 2 of 3 patients enrolled experienced grade 3 edema. Rash is a common AE associated with trametinib and limited patients’ ability to stay on the study, even at the lower dose of 1.0 mg daily. Finally, grade 3 pneumonitis was observed in one patient, raising significant safety concerns for continuing to study this combination. These findings suggest that combined MET and MEK inhibition may potentiate adverse events, including pneumonitis (33%, N=1), edema (67%, N=2), and rash (100%, N=3). Gastrointestinal toxicities, although common with both drugs, were grade 1. Chest pain was reported as a serious adverse event (SAE); however, this patient’s presentation was thought to be unrelated to the study drugs. These findings indicate that the combination of capmatinib and trametinib at the dose levels tested was not well tolerated in this patient population, which is similar to binimetinib and crizotinib combination in a phase I trial in colorectal cancer (15). Overall, this suggests that MET plus MEK inhibitor combinations should be investigated with extreme caution. The lack of radiographic objective responses also suggests limited clinical benefit of this combination. The limitations of this trial included the small sample size and early trial closure, precluding further dose evaluation or expansion and full assessment of efficacy.

In summary, while co-targeting MET and MEK remains a biologically rational strategy to overcome resistance to MET-targeted therapy, the combination of capmatinib and trametinib is poorly tolerated, with all patients discontinuing due to adverse events including grade 3 edema (N=2) and grade 3 pneumonitis (N=1). Further investigation of this combination is not recommended.

## Data Availability

All data produced in the present work are contained in the manuscript.

