## Supplementary Appendix for "Brief Report-Combination Capmatinib and Trametinib in Metastatic MET-driven Non-Small Cell Lung Cancer"

**Detailed Methods**

The study was approved by institutional review boards, and all patients provided written informed consent. Eligible patients were aged 18 years or older and had histologically or cytologically confirmed Stage IV NSCLC with a documented MET exon 14 skipping mutation, MET amplification, or MET fusion, as determined by CLIA-approved assay, and evidence of disease progression per RECIST 1.1 criteria after treatment with at least one prior MET inhibitor (capmatinib, crizotinib, savolitinib, or tepotinib). Patients with other known actionable genomic alterations, prior intolerance to study drugs, interstitial lung disease or pneumonitis, clinically significant heart disease, or prior systemic treatment within the last three weeks were excluded.

Key secondary endpoints included efficacy, as assessed by RECIST 1.1 criteria. Adverse events were assessed by CTCAE v5 criteria. Cycle length was defined as 28 days. Dose-limiting toxicities (DLTs) were defined as any treatment-related toxicity observed within the first cycle of treatment as grade 3 or 4 clinically evident non-hematologic toxicity; grade 4 neutropenia or thrombocytopenia lasting > 7 days or grade 3 or 4 febrile neutropenia or thrombocytopenia with clinically significant bleeding. If No DLT was observed in the initial 3 subjects, enrollment proceeded to the next dose level. If a DLT occurred, three additional subjects were enrolled at the same dose level, for a total of 6. Dose escalation continued until a maximum tolerated dose (MTD) or the maximum administered dose was reached, based on emerging data. If two or more DLTs occurred at a dose level, de-escalation proceeded; if this occurred at dose level -2, the study stopped for lack of tolerability. A minimum of 6 and maximum of 18 patients were planned to be enrolled in this phase I study. Safety and efficacy outcomes included all patients who received at least one dose of capmatinib and trametinib. The final safety analysis was performed after enrollment of three patients.

Assessment of tumor response was performed at baseline and every two cycles beginning Cycle 3 Day 1 for patients whose disease had not progressed since entering the study. Brain imaging was completed at the discretion of the investigator for participants without brain metastases at baseline.

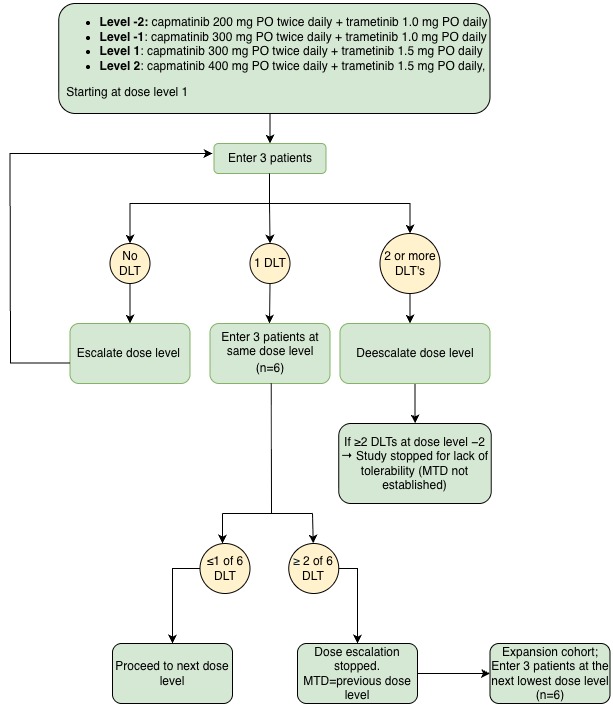

**Supplementary Figure 1:** Study Scheme and dose escalation design. DLT, dose-limiting toxicity; MTD, maximum tolerated dose.

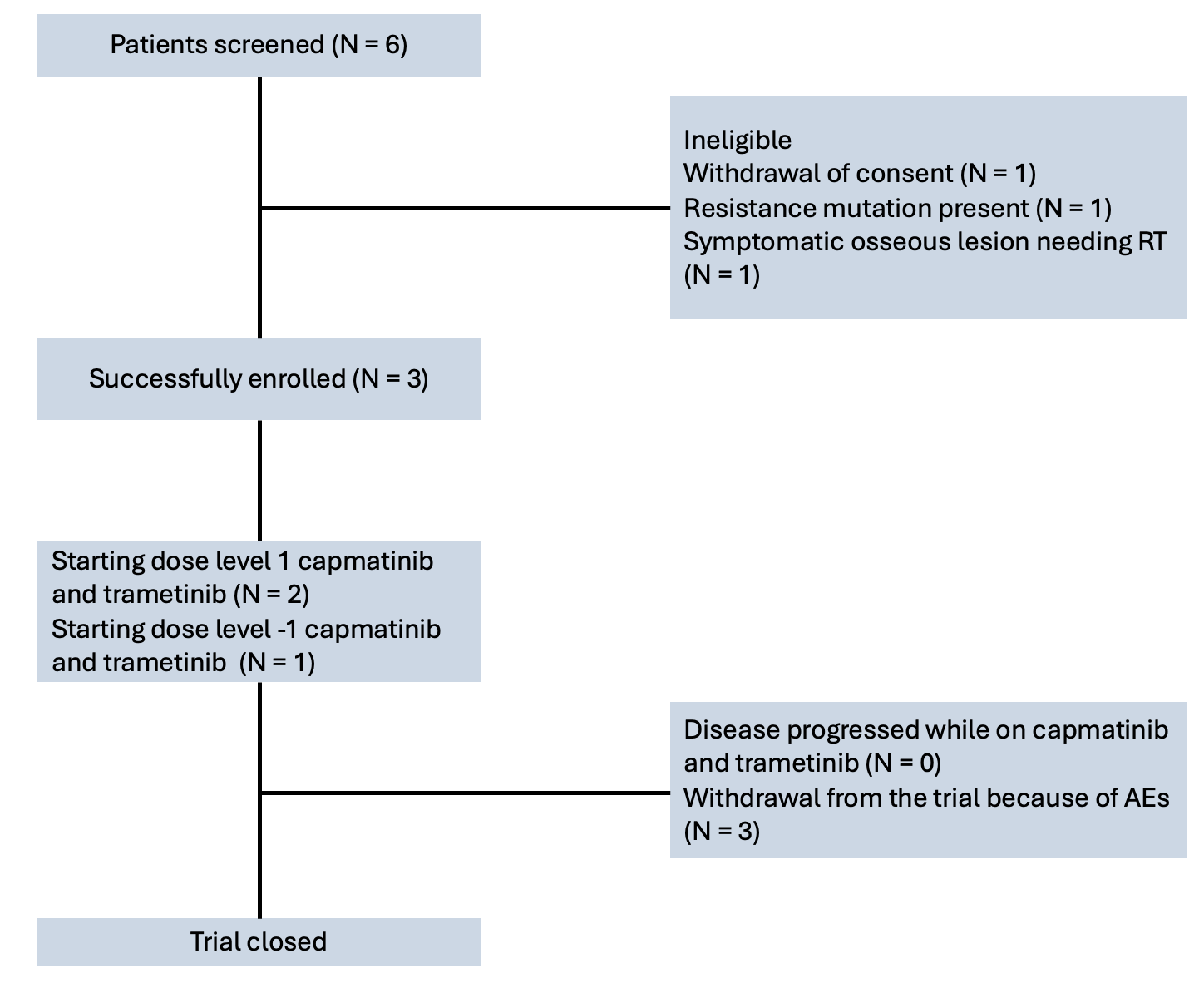

**Supplementary Figure 2:** CONSORT Diagram for patient disposition. AE, adverse event; RT, radiation therapy.

**Supplementary Table 1: Treatment-Related Adverse Events**

| Drug-Related Adverse Event | Total, Any grade No. (%) | Grade 3 -4 No. (%) | Related-Capmatinib | Related-Trametinib | Treatment-related SAE No. (%) | DLT |
| --- | --- | --- | --- | --- | --- | --- |
| Rash | 3 (100%) | 0 (0%) | Unlikely | Definitely | 0 | Yes |
| Rhinorrhea | 1 (33%) | 0 (0%) | Unlikely | Possibly | 0 | No |
| Diarrhea | 1 (33%) | 0 (0%) | Unlikely | Probably | 0 | No |
| Vitreous Detachment | 1 (33%) | 0 (0%) | Unlikely | Possibly | 0 | No |
| Mucositis | 1 (33%) | 0 (0%) | Unlikely | Probably | 0 | No |
| Peripheral edema | 2 (67%) | 2 (67%) | Definitely | Possibly | 0 | Yes |
| Nausea | 1 (33%) | 0 (0%) | Possibly | Possibly | 0 | No |
| Vomiting | 1 (33%) | 0 (0%) | Possibly | Possibly | 0 | No |
| Ocular irritation | 2 (67%) | 0 (0%) | Possibly | Definitely | 0 | No |
| Dyspnea | 2 (67%) | 0 (0%) | Probably | Probably | 0 | No |
| Esophagitis | 1 (33%) | 0 (0%) | Unrelated | Possibly | 0 | No |
| Pneumonitis | 1 (33%) | 1 (33%) | Possibly | Possibly | 1 (33%) | No |
